# Two-way remote monitoring allows effective and realistic provision of home-NIV to COPD patients with persistent hypercapnia

**DOI:** 10.1101/2020.11.08.20227892

**Authors:** Grace McDowell, Maksymilian Sumowski, Hannah Toellner, Sophia Karok, Ciara O’Dwyer, Jamie Hornsby, David Lowe, Chris Carlin

**Author notes:** **Corresponding author:** Dr Chris Carlin.

## Abstract

**Background:** Outcomes for chronic obstructive pulmonary disease (COPD) patients with persistent hypercapnic respiratory failure are improved by long-term home non-invasive ventilation (NIV). Provision of home-NIV presents clinical and service challenges. The aim of this study was to assess outcomes of home-NIV in hypercapnic COPD patients managed remotely.

**Methods:** Retrospective analysis of a dataset of 46 COPD patients with persistent hypercapnic respiratory failure who commenced home-NIV managed by two-way remote monitoring (Lumis, AirView, ResMed) between February 2017 and January 2018. The primary outcome of this study was time to readmission or death at 12 months in patients receiving home-NIV versus a retrospectively identified control cohort of 27 patients with hypercapnic COPD who had not been referred for home-NIV.

**Results:** The median time to readmission or death was significantly prolonged in patients who commenced home-NIV (median 160 days, 95% CI 69.38-250.63) versus the control cohort (66 days, 95% CI 21.9-110.1; p<0.01). Average time to hospital readmission was 221 days (95% CI, 47.77-394.23) and 70 days (95% CI, 55.31-84.69; p<0.05), respectively. Median decrease in bicarbonate level of 4.9mmol/L (p<0.0151) and daytime PCO_2_ 2.2kPa (p<0.032) demonstrate efficacy of home-NIV. A median reduction of 14 occupied bed days per annum versus previous year prior to NIV was observed per patient who continued home-NIV throughout the study period (N=32).

**Conclusion:** These findings confirm the benefits of home-NIV in clinical practice and support the use of two-way remote monitoring as a feasible solution to managing the delivery of home-NIV for COPD patients with persistent hypercapnia.

**KEY MESSAGES:** *What is the key question?:* Do COPD patients with persistent hypercapnia benefit from remotely monitored home-NIV?

*What is the bottom line?:* Home-NIV supported with two-way remote monitoring prolonged time to readmission or death within 12 months in patients with hypercapnic COPD.

*Why read on?:* This is the first study to show that two-way remote monitoring can be an effective and realistic solution in providing access to home-NIV for COPD patients at the necessary scale. The COVID-19 pandemic has mandated remote-management for respiratory care where feasible. This study demonstrates feasibility, safety and efficacy of a remote-management service model for COPD-NIV and provides playbook for adoption by other clinical teams.

## INTRODUCTION

COPD is the second most common cause of emergency hospital admission in the UK, accounting for over 1 million bed days at a cost to the NHS of over £800 million a year ^(1)^. Around a third of those admitted to hospital following an exacerbation of COPD are readmitted within 90 days, which is also strongly associated with post-discharge mortality ^(2)^. Avoiding COPD exacerbations and hospitalisations is noted to be a key priority by COPD patients ^(3, 4)^, and targeting a reduction in these is necessary to address the substantial health and economic burden imposed by COPD.

The risk of hospital readmissions and further life-threatening events is particularly high among patients with a severe exacerbation of COPD that leads to hypercapnic respiratory failure ^(5)^. The first-line treatment for these patients in the acute setting is non-invasive ventilation (NIV) ^(3)^, which has been shown to prevent intubation and invasive mechanical ventilation and reduce hospital mortality ^(6, 7)^. However, it was previously reported that more than 75% of patients treated with NIV for acute hypercapnic respiratory failure were readmitted and nearly 50% died within the first year after discharge ^(5)^.

A growing area of interest to improve outcomes for patients with severe COPD focuses on the application of long-term NIV in the home setting. In a recent landmark study, Murphy et al. ^(8)^ showed that the addition of home-NIV to long-term home oxygen therapy in patients that remained severely hypercapnic 2 to 4 weeks after an exacerbation delayed and reduced hospital readmissions at 12 months. A benefit on 12-month overall survival was noted in an earlier randomised controlled trial involving stable hypercapnic patients treated with home-NIV ^(9)^. The driver of clinical improvements across both studies can be attributed to higher inspiratory pressures targeting a reduction in CO_2_ in patients who were persistently hypercapnic. A task force of the European Respiratory Society has since adopted home NIV as recommended treatment for COPD patients presenting with persistent hypercapnic respiratory failure ^(10)^.

Overall, the existing body of research suggests that there are some open questions with regards to patient selection and timing of home-NIV ^(11, 12)^. For example, only 5% of patients screened in HOT-HMV study were recruited to the trial, raising questions about the external validity of NIV randomised controlled trial (RCT) results. Many of the patients excluded from NIV RCTs meet current guidance critieria for home NIV provision. Establishing whether beneficial outcomes from home NIV COPD RCTs can be matched with routine clinical adoption is required.

The feasibility of delivering home-NIV to patients outside of controlled clinical trial settings also remains to be established, particularly in the context of COVID-19 pandemic. Provision for elective inpatient NIV initiation and titration beyond clinical trials is limited, patients generally wish to avoid hospitalisations and severity of their illness limits capacity for outpatient attendances. Regular follow-up helps to monitor the effectiveness of ventilation, encourages treatment adherence and optimises patient comfort and ventilator settings, but realistic delivery of intensive follow-up is problematic ^(13)^. The COVID-19 pandemic has presented additional challenges to home NIV provision. Overall healthcare service pressures, social distancing requirements including need to protect vulnerable patients from nosocomial COVID-19, and infection control requirements for clinicians, with NIV classified as an aerosol generating procedure will all continue to impact on breathing support service capacity.

It has been demonstrated that COPD patients at high risk for exacerbations can be taught to self-manage when offered ongoing support ^(14, 15)^. Early evidence that compares remotely monitored COPD patients with usual face-to-face care is encouraging in terms of patient quality of life and number of hospital admissions ^(16)^. With the recent advent of two-way tele-monitoring, healthcare providers can view live NIV data from patients, adjust ventilator settings remotely and facilitate personalised care. A combination of patient education, self-management and remote monitoring may therefore be a realistic support pathway for home-NIV. Using this to channel shift service provision, replacing some aspects of inpatient NIV setup and/or face-face outpatient NIV follow-up on an individualised basis is attractive, particularly to mitigate risks and bolster service provision in face of COVID-19 related challenges.

NHS Greater Glasgow and Clyde (GG&C) implemented a remote-monitored home-NIV model for its COPD population in 2017. The present study retrospectively analysed all patients who were commenced on therapy over the first 12 months of this service, with aim of determining whether outcomes similar to RCTs were achieved in a real-world cohort of hypercapnic COPD patients with typical comorbidities (which would have excluded many from NIV RCTs) who are managed with remote-monitored home NIV. The primary outcome was median time to readmission or death over 12 months in patients receiving home-NIV versus a control cohort.

## METHODS

### Study design and patients

This study is part of programme of work analysing outcomes in a dataset of COPD patients provided by NHS GG&C Safe Haven. Local Privacy Advisory Committee approval was obtained for release of de-identified data for this study.

Two cohorts were sampled from the database as outlined in Figure 1. The home-NIV cohort consisted of 42 consecutive patients with COPD who commenced home-NIV between February 25^th^ 2017 and January 25^th^ 2018 at the Queen Elizabeth University Hospital. COPD diagnosis was confirmed as per GOLD guidelines, and was the primary diagnosis responsible for hypercapnic respiratory failure in all patients in this cohort. Hypercapnic respiratory failure was defined as PCO^2^ >7kPa at least 2 weeks after index acute exacerbation and/or presence of persisting hypercapnia across current and previous COPD episodes, with deferred NIV assessment for attempted follow up post episode judged inappropriate. 28 patients in this cohort continued home-NIV throughout the 12-month study period (‘NIV users’). 14 patients discontinued home-NIV due to poor acceptance despite individualised interventions to optimise therapy during the study period (‘NIV non-users’).

**Figure 1.**
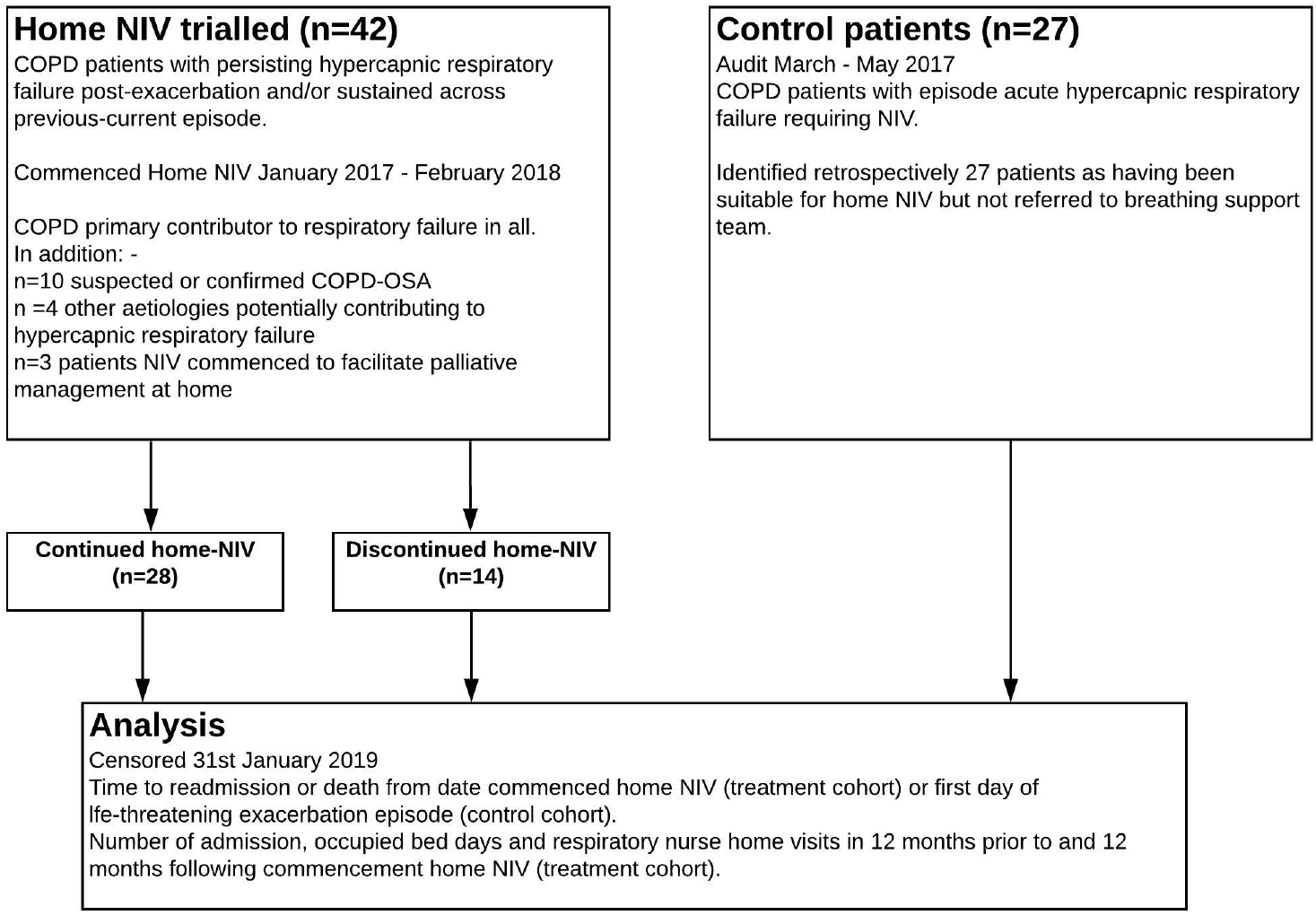
Study participant flow diagram

The control cohort comprised 27 patients treated with acute NIV at the Queen Elizabeth University Hospital between March and November 2017 following a life-threatening exacerbation of COPD that resulted in hypercapnic respiratory failure. This was in the period prior to adoption of routine screening of all acute NIV patients for home NIV at our institution. Retrospective review noted that patients in this group were suitable for home-NIV but they were not referred to the home-NIV service during the follow up period of this study. None of these patients ‘crossed over’ to commence home NIV during the study’s observed follow up period.

All patients were noted to be receiving guideline-based COPD care, including home oxygen therapy unless contraindicated.

### Intervention

Since early 2017 our practice has been to offer trialling home NIV to COPD patients with persistent hypercapnia (PcCO2 >7kPa) at stable status, or during an acute episode if there has been recurrent hypercapnic respiratory failure where deferring commencing home NIV to outpatient review is judged impractical or unsafe by patient-clinician consensus. Often this decision to offer home NIV within an acute episode is informed by high serum bicarbonate levels (implying chronic hypercapnia) and/or presence of suspected or confirmed OSA overlapping with severe COPD.

Patients in the home-NIV cohort were commenced on remote monitored home-NIV in iVAPS auto-EPAP mode (Lumis 150 ST-A, AirView, ResMed) if persisting hypercapnia was present at day case review 2 to 4 weeks following hospital discharge (n=14/42) or during the index hospital admission if persisting hypercapnia had been demonstrated across previous COPD episodes as per above noted criteria (n=28/42)

Patients initiated on home-NIV consented to their data being accessed and shared on the AirView platform by the necessary healthcare professionals. AirView data review was used to inform routine clinical care and identify NIV therapy issues (usage, leak) as well as ventilation data patterns supporting optimized NIV provision. Patient telephone, community or clinic follow up was individualized based on and informed by the remote monitoring data. AirView platform was used to make NIV device therapy changes, when indicated.

The Supplementary Material provides an overview of the COPD NIV therapy protocol implemented at NHS GG&C, with typical follow-up schedule as well as representative remote monitoring data. Remote-monitoring pathway is used to support daycase NIV initiation (rather than elective hospital admission), early hospital discharge (if NIV initiated during inpatient episode our institution) or inpatient NIV initiation at another hospital (rather than hospital-hospital transfer). Remote-monitoring data is reviewed at day 1-2, day 5-7 and weekly thereafter, combined where required with telephone or video consultation, until treatment is optimized. Remote-adjustments to iVAPS-autoEPAP NIV settings, adjustment to NIV interface and face-face at home or daycase review arrangement are made where remote-monitoring and consultation data indicates requirement. Stability is judged based on patient comfort and symptoms, acceptable NIV usage durations, minimized unintentional leak and appropriate pressure support and other ventilator parameters. Clinic follow up within 8-12 weeks including repeat capillary blood gases is scheduled for stable patients who can attend. Patients who are having persisting difficulties establishing home NIV despite remote-monitoring inputs are offered elective admission.

### Outcome measures

Baseline descriptive data were recorded including gender, age, BMI, predicted FEV1% as well as comorbidities that could potentially contribute to hypercapnia. The primary outcome was time to readmission or death, censored at date of admission, date of death or 25^th^ January 2019. Secondary outcome measures included time to hospital admissions and overall survival in the home-NIV and control cohort.

Subgroup analyses of the home-NIV cohort explored differences between NIV users and NIV non-users in the primary and secondary outcome measures. Changes in healthcare usage (number of hospital admissions, occupied bed days (OBDs) and respiratory nurse home visits) were evaluated in NIV users and NIV non-users before and after home-NIV. Changes in capillary blood gases were analysed in the home-NIV cohort in the form of capillary blood gas PCO_2_ and bicarbonate (where available).

### Statistical analyses

Baseline characteristics of the study population are presented as mean (standard deviation), median (interquartile range) or count (percentage), as appropriate. Primary and secondary study outcome measures were compared between the home-NIV and control cohort using Kaplan-Meier survival analysis and the Mantel-Cox log rank test.

Additional subgroup analyses compared primary and secondary outcome measures between the NIV user group and the NIV non-user group alongside the control cohort using Kaplan-Meier and Mantel-Cox tests. Changes in healthcare usage (number of hospital admissions, OBDs, and respiratory nurse home visits) and capillary blood gas PCO_2_ and bicarbonate between NIV users, NIV non-users and the control cohort were analysed using Wilcoxon signed-rank test. Statistical analyses were conducted using IBM SPSS Statistics V.24 (IBM, New York, USA) and GraphPad Prism v7.0 (GraphPad Software, San Diego, USA).

### Patient and Public Involvement

Due to the nature of a retrospective analysis, the research was undertaken without patient involvement. Patients were not invited to comment on the study design and were not consulted to develop patient relevant outcomes, interpret the results or to contribute to the writing or editing of this document for readability or accuracy.

## RESULTS

### Baseline characteristics

Baseline characteristics are shown in Table 1. Gender, BMI and age are similar across all study cohorts except for a higher rate of males, lower BMI and a lower rate of notable comorbidities (suspected or confirmed overlapping OSA, long term opiate therapy) in the NIV non-user subgroup. The FEV1% predicted value was around 40% across all study groups, in line with a “severe” classification of COPD ^(3)^ particularly as in many patients the spirometry was an historical rather than contemporary result.

**Table 1.** Baseline characteristics

|  | Total |  | Control cohort |  | Home-NIV |  |  |  |  |  |
| --- | --- | --- | --- | --- | --- | --- | --- | --- | --- | --- |
|  |  | *n* |  | *n* | *Total* | *n* | *NIV users* | *n* | *NIV non-users* | *n* |
| Gender, n (% female) | 49 M 20F<br>(28.9) | 69 | 18M 9F<br>(29.6) | 27 | 30M 12F<br>(28.5) | 42 | 17M 11F<br>(39.3) | 28 | 13M 1F (7.1) | 14 |
| Mean age (SD) | 64.55 (8.74) | 69 | 66.48 (8.8) | 27 | 63.31 (8.58) | 42 | 62.54 (8.75) | 28 | 64.86 (8.31) | 14 |
| Mean BMI (SD) | 27.6 (9.09) | 53 | 26.02 (7.06) | 27 | 29.23 (10.71) | 26 | 32.23 (10.63) | 16 | 21.41 (9.42) | 10 |
| FEV1% Predicted Mean (SD) | 40.63 (16.63) | 52 | 41.29 (18.61) | 21 | 40.19 (15.45) | 31 | 39.63 (16.28) | 19 | 41.08 (14.7) | 12 |
| Comorbidity potentially contributing to hypercapnia, n (%) | - | - | - | - | 14 (37.8) | 37 | 11 (44) | 25 | 3 (21.4) | 12 |

### Time to readmission or death

The median time to readmission or death was significantly prolonged in patients with persisting hypercapnic respiratory failure treated with home-NIV compared to a control cohort of hypercapnic COPD patients (*p<*0.01, see Figure 2a). Subgroup analyses showed significant differences between the NIV user subgroup versus both the NIV non-user group and the control cohort (both *p*<0.001). Improvement in time to readmission or death was not achieved in patients who discontinued home-NIV (Figure 2b). Table 2 summarises time to readmission or death for each group.

**Table 2.**
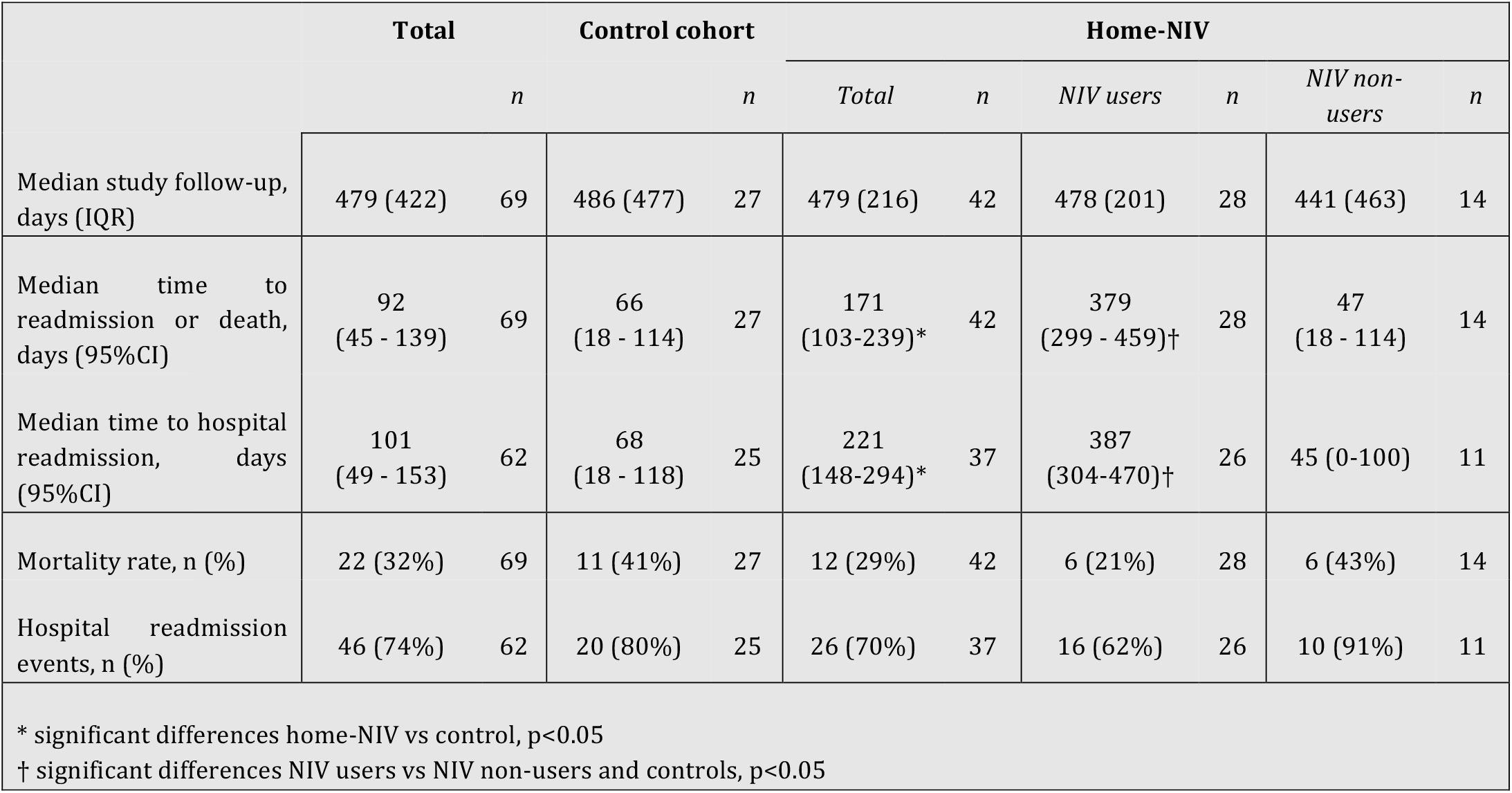
Primary and secondary study endpoints per study population

**Figure 2.**
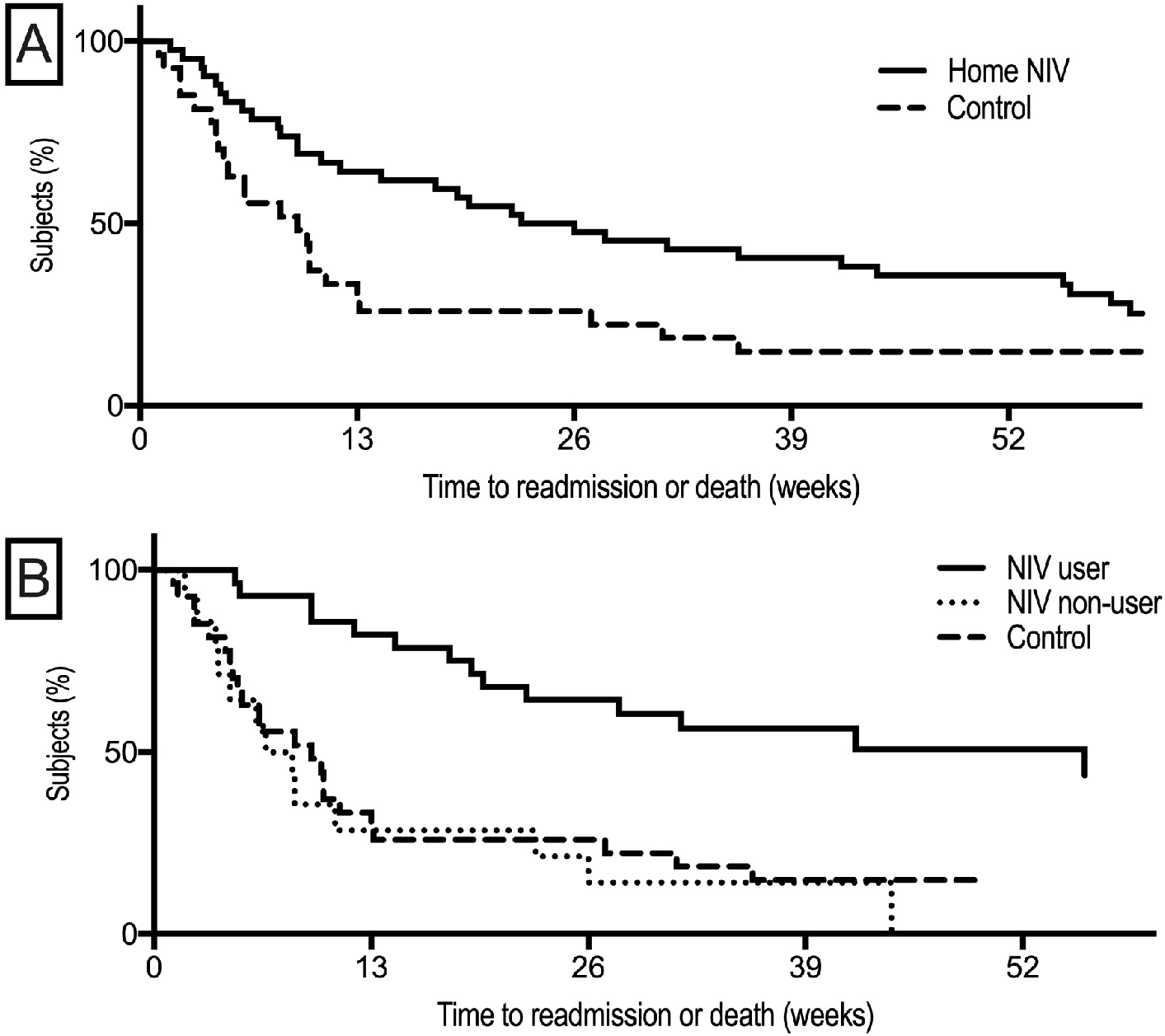
Kaplan-Meier plot of time to readmission or death from study initiation to the end of study follow-up. (A) Primary analysis shows significant differences between the home-NIV and the control cohort. (B) Subgroup analyses showed that the improvement in 12-month readmission avoidance is noted only in patients who continue home-NIV throughout the study period.

### Median number of days to hospital readmission

Time to hospital readmission followed the same pattern as time to readmission or death. Median time to hospital readmission was 221 days for the home-NIV cohort (95% CI, 148 - 294) and 68 days (95% CI, 18 N - 118; *p*<0.05) for the control cohort. Subgroup analyses showed that time to hospital readmission was significantly improved in NIV-users when compared to NIV non-users and the control group (both *p*<0.01). There was no significant difference comparing the control group and NIV non-users (*p*=0.38).

### Overall survival

12-month overall survival was 78.6% in the home-NIV cohort and 59.3% in the control cohort. Patients that continued to use home-NIV during the study period had a 12-month overall survival rate of 85.7%. Due to the low number of recorded mortality events, group differences were not statistically significant in the primary (*p*=0.066) or subgroup analyses (*p*=0.07).

### Healthcare usage

Service usage in NHS GG&C by the home-NIV cohort in the 12 months prior to commencing home-NIV (Pre-NIV) and the 12 months following initiation of home-NIV (Post-NIV) are outlined in Table 3. A significant reduction in total number of admissions and OBDs is noted following initiation of home-NIV across all patients in the home-NIV cohort, but is particularly pronounced in NIV users (*p*<0.001, Figure 3). The data equate to a median reduction of 14 OBDs per annum per patient who continued remote-monitored home-NIV. Requirements for respiratory nurse home visits did not change significantly with the initiation of home-NIV.

**Table 3.** Changes in healthcare usage before and after home-NIV

|  | Pre-NIV | Post-NIV | n | p |
| --- | --- | --- | --- | --- |
| **Number of hospital admissions** | 127 | 67 | 42 | 0.0005 |
| *NIV users* | 78 | 35 | 28 | 0.0015 |
| *NIV non-users* | 49 | 32 | 14 | 0.1477 |
| **Occupied bed days (OBDs)** | 1131 | 414 | 42 | <0.0001 |
| *NIV users* | 775 | 230 | 28 | 0.0005 |
| *NIV non-users* | 356 | 184 | 14 | 0.0254 |
| **Respiratory nurse home visits** | 69 | 67 | 24 | 0.9158 |
| *NIV users* | 41 | 43 | 14 | 0.7134 |
| *NIV non-users* | 28 | 24 | 10 | 0.8711 |

**Figure 3.**
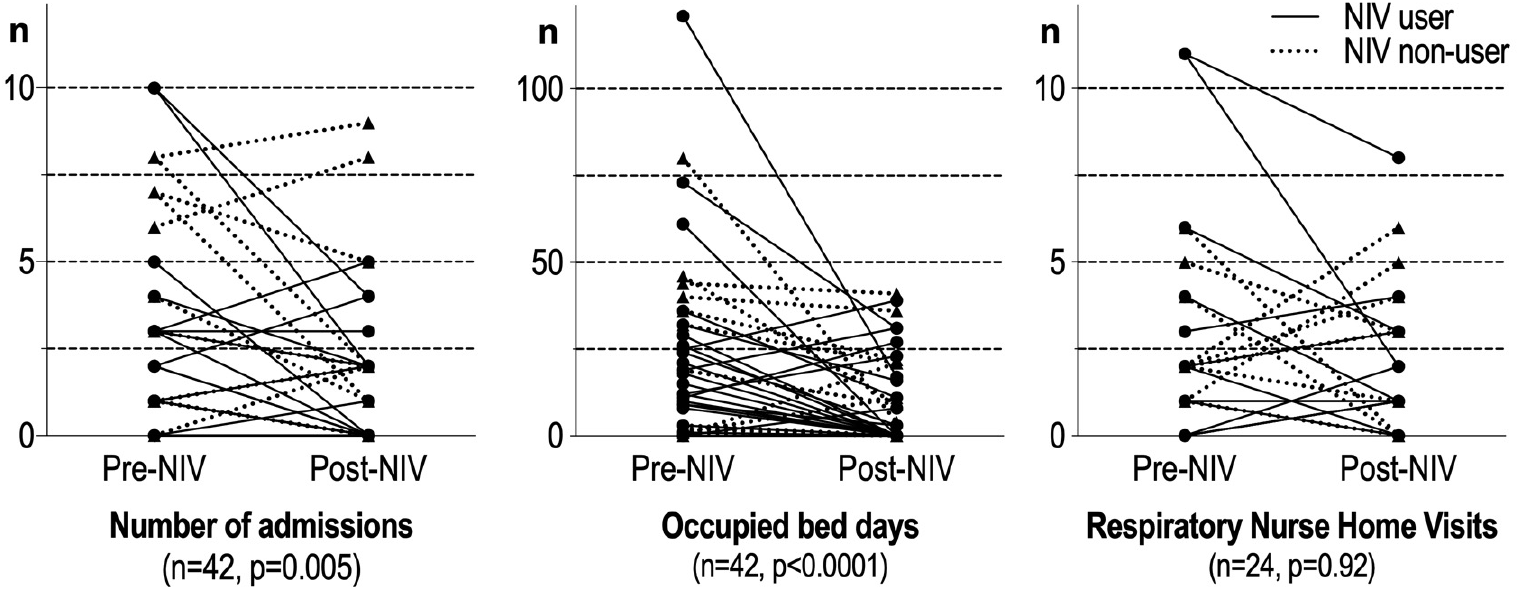
Changes in healthcare usage before and after home-NIV for NIV users (circle) and NIV non-users (triangle). Data on respiratory nurse home visits was not available in electronic health records for the 18 patients whose residence is outside our health board.

### Blood gas exchange

Capillary blood gas measurements were available in 21 patients before and after home-NIV. Significant improvements in median PCO_2_ (2.2kPa, p<0.05) and bicarbonate (4.9mmol/L, p<0.05) measured at follow-up after initiation of home-NIV relative to measurements at baseline indicate control of hypercapnic respiratory failure (Figure 4).

**Figure 4.**
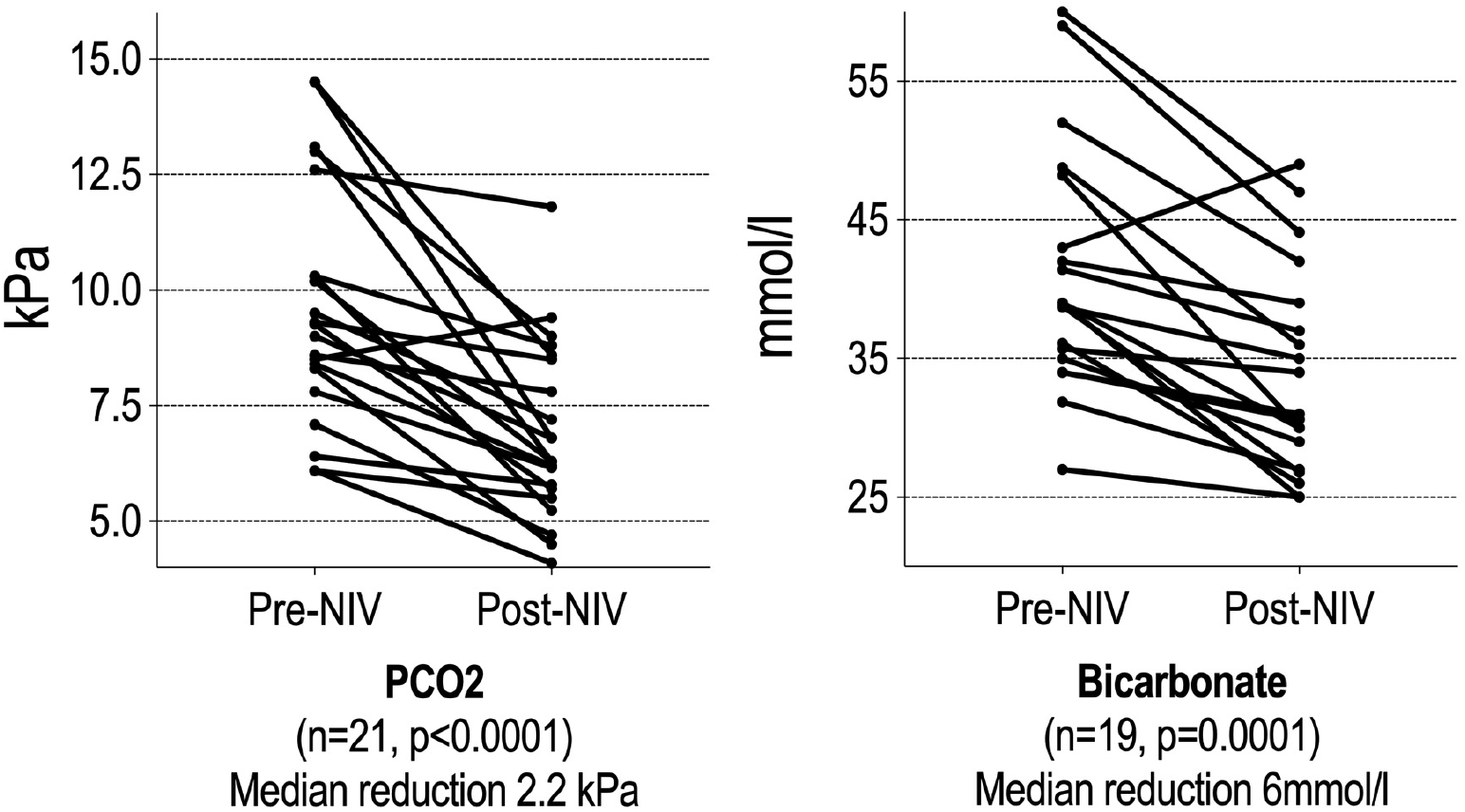
Changes in blood gas measurements at baseline and follow up after home-NIV initiation in NIV users. Data availability limited to subset of patients who attended for face-face follow up and had some or all components of post NIV blood gas results inputted into electronic health record (including 2 patients who had PCO2 but not bicarbonate result available). 2 patients had PCO2 <7kPa at time of NIV initiation but had other standard indications to commence home NIV.

## DISCUSSION

This study confirms the benefits of home-NIV in COPD patients with persisting hypercapnic respiratory failure in clinical practice. Patients in the home-NIV cohort had significantly fewer hospital readmissions compared to the control cohort, with the greatest improvements seen in those who continued with home-NIV throughout the 12-month observed follow-up period. Moreover, these data support the use of two-way remote monitoring as a feasible solution to managing the delivery of home-NIV, maintaining care-quality while also substantially reducing demand on healthcare resources.

The current results are consistent with the HOT-HMV trial by Murphy et al. ^(8)^ reporting delayed and reduced hospital readmissions in COPD patients randomised to receive home-NIV compared to patients treated with home oxygen therapy alone. While there are some important differences to consider between the HOT-HMV trial and this study - less severe documented airflow obstruction and higher BMI in this study’s patient cohort - reporting similar outcomes to the HOT-HMV trial is encouraging. Our data indicates that home-NIV is effective in a typical population of patients presenting in routine clinical practice with phenotypes which would have potentially excluded them from RCT inclusion. The secondary analyses also broadly support the interpretation of HOT-HMV and other landmark COPD home-NIV trials ^(9, 17)^ indicating that patient selection for home-NIV should be based on persisting hypercapnic respiratory failure, and that optimising NIV to target improvement in hypercapnia is appropriate. The comparable outcomes of this study and HOT-HMV importantly provide reassurance about safety and quality of a COPD home-NIV service model utilising assistive two-way remote-monitoring technology.

In the HOT-HMV trial, home-NIV was noted to reduce exacerbation-related costs (by £1,141 per case) and patient-reported costs (by £2,337) relative to the control arm. NIV device cost and cost per physician visit had the greatest impact on cost per QALY ^(18)^. In line with this, the present study notes a considerable reduction in healthcare usage among home-NIV users. In addition to significant decreases in hospital admissions, a median reduction of 14 occupied bed days per annum was observed per patient who continued home-NIV. It seems likely that a COPD home-NIV management strategy based on remote-monitoring and individualised follow-up will reduces physician visit and patient travel costs and impact positively on patient quality of life. Remote-monitoring based service model should reduce home NIV costs and potentially further improve the quality of life benefit. Future assessments are required to expand on the cost-effectiveness of home-NIV and a proactive COPD service model based on remote-monitoring.

Among various patient groups using NIV at home, remote monitoring has been found to be non-inferior and at times more effective than usual face-to-face support, preferred by patients and associated with reduced healthcare utilisation ^(16, 19-21)^. The additional channel of two-way patient engagement – that is, early intervention with an NIV therapy change to optimise settings based on remote-monitoring data – may prove particularly valuable to ensure continued treatment adherence ^(22)^. Monitoring patients is a prerequisite for successful continuation of NIV at home. Maintaining the required level of contact face-to-face is particularly challenging for severe COPD patients, who are often not fit to travel or who may require immediate intervention. Remote-monitoring data can be utilised to support and enhance routine clinical care allowing positive endorsement to be relayed when monitoring data is reassuring, prioritising and focusing patient-clinician interactions when issues are noted. We show for the first time that initiation and follow-up based exclusively on NIV two-way remote monitoring can be an effective and realistic solution to providing access to home-NIV to severe COPD patients at the necessary scale. Benchmarking of outcomes, with similar findings to published RCT data provides reassurance about safety and maintained care-quality with a remote-monitoring based approach to home NIV for hypercapnic COPD. Our findings broadly complement those from recent study reported from the Netherlands, which demonstrated cost-effective provision of home initiation of NIV for stable hypercapnic COPD patients utilising remote-monitoring of ventilator and transcutaneous CO2 data ^(23)^. As key additions, our data suggest that remote-monitoring can be used to safely support patients with persisting hypercapnia who are commenced on home NIV at an acute episode as well as at stable status, and that overnight transcutaneous CO_2_ monitoring can be omitted from routine follow-up of COPD patients on home-NIV. Undertaking routine transcutaneous monitoring in this patient cohort would not be realistic in routine clinical practice at scale. Continued supervision of this approach with reporting of outcomes to ensure safety and quality of home-NIV therapy, alongside continued evaluation of other endpoints for respiratory failure monitoring is required.

Our approach to home-NIV setup for hypercapnic COPD patients differs from published protocols. We have accrued considerable experience with volume assured pressure support NIV modes at our centre. Our experience is that these allows NIV optimisation to be undertaken more efficiently with enhanced patient comfort and improved treatment quality as well as additional benefits including anticipation that treatment will be responsive to predictable fluctuations in a patient’s condition. We commence NIV treatment for COPD patients in iVAPS autoEPAP mode, targeting symptom benefit, remote-monitored ventilation patterns and follow-up capillary blood gas results (see Supplementary data file), rather than in-hospital titration of NIV in ST mode, targeting high pressure support with transcutaneous CO^2^ monitoring. Satisfactory clinical outcomes and follow-up blood gas data (median reduction PCO2 2.2 kPa) in this study provide reassurance about the quality and safety of this NIV setup protocol. Clinical user experience with this approach is positive, and it achieves reduction in occupied bed days with no additional staffing support required to deliver the service. Whether matched clinical outcomes and similar efficiency would be achieved with remote-monitored NIV utilising ST mode (potentially with at home titration) requires additional study.

Our subgroup analyses consistently showed that outcomes in patients who discontinue home-NIV align closely with outcomes from the control cohort. This suggests that patients with severe hypercapnic COPD are not negatively affected by the process of initiating home-NIV. Further exploration is indicated in this sub-population of patients who do not tolerate long-term NIV at home. It could be that greater attention to patient activation, treatment provision, ventilation optimisation or level of contact is required. The patients who discontinued home NIV had significant remote-management based scrutiny and input, and were offered ‘standard’ in-hospital and/or outpatient face-face attendance to try and maintain NIV use. Other centres have noted progressive improvements in home NIV adherence rates with multi-disciplinary ‘NIV failure’ clinics^25^. Adoption of remote-monitoring can improve the workflow and prioritisation of NIV MDT activity. Whether additional MDT input than that provided in our described model would improve long term NIV adherence is uncertain. The possibility that there are different responder groups regardless of optimisation efforts should also be considered. In our cohort, there was a higher proportion of female patients and higher mean BMI in NIV users. We can speculate that these differences might reflect home support or early symptom benefit differences from home NIV: obese patients may be more likely to have OSA overlapping with hypercapnic COPD. Characterisation of these and other factors with remote-monitoring data comparing sustained users and non-users in future studies may allow further evaluation to a service model with evidence-based proactive prioritisation: intensive focused MDT input to those patients who require it, and minimised input to those where it is unnecessary or will be non-contributory. Lastly, the presence of a control cohort, which consisted of patients who may have benefited but were not referred for home-NIV from within an active tertiary NIV centre, highlights the need for clinician education and other efforts to ensure equitable patient access to this evidence-based intervention.

This study had several strengths, including the use of clinically meaningful outcomes and the real-world nature of the patient cohorts. However, we acknowledge several limitations. Treatment allocation was not randomised and the impact of unrecognised confounding factors cannot be ruled out. We also did not have complete data on demographics, comorbidities or provision of and adherence to other COPD treatments to ensure cohorts were otherwise matched. The statistical analyses of some of the subgroup analyses should be considered exploratory due to the limited sample size and the potential issue of multiple testing. Finally, this study was not powered to find a difference in survival. While a survival benefit of home-NIV has been previously demonstrated in a similar patient population ^(9)^, clear evidence of improved survival is still lacking and should be investigated in larger prospective trials.

## CONCLUSION

COPD patients with persistent hypercapnic respiratory failure have poor outcomes with limited treatment options available. To our knowledge, this is the first study to confirm the benefits of remote-managed home-NIV in this group of COPD patients as they typically present in clinical practice. Home-NIV prolonged the time to readmission or death within 12 months in patients with persistent hypercapnia following an acute exacerbation of COPD. In addition to being the outcomes that COPD patients rate as most important ^(24)^, exacerbation and hospitalisation avoidance address the substantial economic burden imposed by COPD. We report significant reductions in healthcare usage among home-NIV users and demonstrate that two-way remote monitoring can be an effective and realistic solution to providing access to home-NIV for hypercapnic COPD patients. The COVID-19 pandemic has presented considerable challenges to home-NIV service provision. Our data provides reassurance that a service model based on outpatient or truncated inpatient NIV initiation and remote-monitoring based follow up allows face-face contact to be safely minimised, reducing COVID-19 transmission risks whilst maintaining NIV care-quality.

## Data Availability

Data in the manuscript is available in NHS GG&C SafeHaven. Data could be provided with standard approval access request.
https://www.nhsggc.org.uk/about-us/professional-support-sites/safe-haven/services/#

## ACKNOWLEDGMENTS

We gratefully acknowledge the comprehensive contribution of the respiratory physiologist and nurse specialist teams in NHS GG&C to the positive outcomes reported in this paper: they have adapted service models to realise benefits from assistive technologies, and continue to be enthusiastically committed to improving patient outcomes and providing realistic medicine.

## REFERENCES

1. NHS. COPD Commissioning Toolkit: A Resource for Commissioners https://assets.publishing.service.gov.uk/government/uploads/system/uploads/attachment_data/file/212876/chronic-obstructive-pulmonary-disease-COPD-commissioning-toolkit.pdf : NHS Medical Directorate; 2012 [updated 2012].

2. Hartl S, Lopez-Campos JL, Pozo-Rodriguez F, Castro-Acosta A, Studnicka M, Kaiser B, et al. Risk of death and readmission of hospital-admitted COPD exacerbations: European COPD Audit. European Respiratory Journal. 2016;47(1):113–21.

3. Vogelmeier CF, Criner GJ, Martinez FJ, Anzueto A, Barnes PJ, Bourbeau J, et al. Global Strategy for the Diagnosis, Management, and Prevention of Chronic Obstructive Lung Disease 2017 Report. GOLD Executive Summary. Am J Respir Crit Care Med. 2017;195(5):557–82.

4. Vestbo J, Hurd SS, Agusti AG, Jones PW, Vogelmeier C, Anzueto A, et al. Global strategy for the diagnosis, management, and prevention of chronic obstructive pulmonary disease: GOLD executive summary. Am J Respir Crit Care Med. 2013;187(4):347–65.

5. Chu CM, Chan VL, Lin AW, Wong IW, Leung WS, Lai CK. Readmission rates and life threatening events in COPD survivors treated with non-invasive ventilation for acute hypercapnic respiratory failure. Thorax. 2004;59(12):1020–5.

6. Kondo Y, Kumasawa J, Kawaguchi A, Seo R, Nango E, Hashimoto S. Effects of non-invasive ventilation in patients with acute respiratory failure excluding post-extubation respiratory failure, cardiogenic pulmonary edema and exacerbation of COPD: a systematic review and meta-analysis. Journal of Anesthesia. 2017;31(5):714–25.

7. Osadnik CR, Tee VS, Carson-Chahhoud KV, Picot J, Wedzicha JA, Smith BJ. Non-invasive ventilation for the management of acute hypercapnic respiratory failure due to exacerbation of chronic obstructive pulmonary disease. Cochrane Database of Systematic Reviews. 2017(7).

8. Murphy PB, Rehal S, Arbane G, Bourke S, Calverley PMA, Crook AM, et al. Effect of home noninvasive ventilation with oxygen therapy vs oxygen therapy alone on hospital readmission or death after an acute COPD exacerbation: A randomized clinical trial. Jama. 2017;317(21):2177–86.

9. Köhnlein T, Windisch W, Köhler D, Drabik A, Geiseler J, Hartl S, et al. Non-invasive positive pressure ventilation for the treatment of severe stable chronic obstructive pulmonary disease: a prospective, multicentre, randomised, controlled clinical trial. The Lancet Respiratory Medicine. 2014;2(9):698–705.

10. Ergan B, Oczkowski S, Rochwerg B, Carlucci A, Chatwin M, Clini E, et al. European Respiratory Society guidelines on long-term home non-invasive ventilation for management of COPD. Eur Respir J. 2019;54(3).

11. Duiverman ML. Noninvasive ventilation in stable hypercapnic COPD: what is the evidence? ERJ open research. 2018;4(2):00012–2018.

12. Storre JH, Callegari J, Magnet FS, Schwarz SB, Duiverman ML, Wijkstra PJ, et al. Home noninvasive ventilatory support for patients with chronic obstructive pulmonary disease: patient selection and perspectives. Int J Chron Obstruct Pulmon Dis. 2018;13:753–60.

13. Arnal JM, Texereau J, Garnero A. Practical Insight to Monitor Home NIV in COPD Patients. COPD. 2017;14(4):401–10.

14. Bucknall CE, Miller G, Lloyd SM, Cleland J, McCluskey S, Cotton M, et al. Glasgow supported self-management trial (GSuST) for patients with moderate to severe COPD: randomised controlled trial. BMJ. 2012;344:e1060.

15. Milligan M, Marshall S, Begum R, Anderson D. Innovative approach to COPD improves disease impact, quality of life and reduces hospital admissions in Glasgow. European Respiratory Journal. 2016;48(suppl 60):PA722.

16. McLean S, Nurmatov U, Liu JL, Pagliari C, Car J, Sheikh A. Telehealthcare for chronic obstructive pulmonary disease. Cochrane Database Syst Rev. 2011(7):Cd007718.

17. Struik FM, Sprooten RT, Kerstjens HA, Bladder G, Zijnen M, Asin J, et al. Nocturnal non-invasive ventilation in COPD patients with prolonged hypercapnia after ventilatory support for acute respiratory failure: a randomised, controlled, parallel-group study. Thorax. 2014;69(9):826–34.

18. Murphy PB, Brueggenjuergen B, Reinhold T, Fusfeld L, Gu Q, Goss T, et al. Cost-Effectiveness of Home Oxygen Therapy-Home Mechanical Ventilation (HOT-HMV) for the Treatment of Chronic Obstructive Pulmonary Disease (COPD) with Chronic Hypercapnic Respiratory Failure Following an Acute Exacerbation of COPD in the United Kingdom (UK). A102 DETERMINANTS OF OUTCOMES AND HIGH-VALUE CARE IN COPD: American Thoraic Society; 2018. p. A2517.

19. Moreira J, Freitas C, Redondo M, Pinto T, Pires F, Sucena M, et al. Compliance with home non-invasive mechanical ventilation in patients with chronic respiratory failure: Telemonitoring versus usual care surveillance - a randomized pilot study. European Respiratory Journal. 2014;44(Suppl 58):P447.

20. Young J, Ashforth N, Price K, Wilson A, Nickol A. Remote monitoring of home non-invasive ventilation: a feasibility study. European Respiratory Journal. 2018;52(suppl 62):PA1669.

21. Pinto A, Almeida JP, Pinto S, Pereira J, Oliveira AG, de Carvalho M. Home telemonitoring of non-invasive ventilation decreases healthcare utilisation in a prospective controlled trial of patients with amyotrophic lateral sclerosis. J Neurol Neurosurg Psychiatry. 2010;81(11):1238–42.

22. Woehrle H, Arzt M, Graml A, Fietze I, Young P, Teschler H, et al. Effect of a patient engagement tool on positive airway pressure adherence: Analysis of a German healthcare provider database. Sleep Med. 2018;41:20–6.

23. Duiverman ML, Vonk JM, Bladder G, Van Melle JP, Nieuwenhuis J, Hazenberg A, et al. Home initiation of chronic non-invasive ventilation in COPD patients with chronic hypercapnic respiratory failure: a randomised controlled trial. Thorax. 2019; doi:10.1136/thoraxjnl-2019-213303.

24. Zhang Y, Morgan RL, Alonso-Coello P, Wiercioch W, Bala MM, Jaeschke RR, et al. A systematic review of how patients value COPD outcomes. Eur Respir J. 2018;52(1).

25. Masoud O, Ramsay M, Suh E-S, Kaltsakas G, Srivastava S, Pattani H, et al. Long term adherence to home mechanical ventilation: a 10-year retrospective, single-centre cohort study. J Thoracic Dis 2020; 12(S2): S120–128

